# Investigation of pooling strategies using clinical COVID-19 samples for more efficient diagnostic testing

**DOI:** 10.1101/2020.08.10.20171819

**Authors:** Samantha H. Adikari, Emily Z. Alipio Lyon, Attelia D. Hollander, Alina Deshpande, Elizabeth Hong-Geller

## Abstract

When testing large numbers of clinical COVID-19 samples for diagnostic purposes, pooling samples together for processing can offer significant reductions in the materials, reagents, time, and labor needed. We have evaluated two different strategies for pooling independent nasopharyngeal swab samples prior to testing with an EUA-approved SARS-CoV-2 RT-qPCR diagnostic assay. First, in the Dilution Study, we assessed the assay's ability to detect a single positive clinical sample diluted in multiple negative samples before the viral RNA extraction stage. We observed that positive samples with Ct values at ~30 can be reliably detected in pools of up to 30 independent samples, and positive samples with Ct values at ~35 can be detected in pools of 5 samples. Second, in the Reloading Study, we assessed the efficacy of reloading QIAamp viral RNA extraction columns numerous times using a single positive sample and multiple negative samples. We determined that one RNA extraction column can be reloaded with up to 20 clinical samples (1 positive and 19 negatives) sequentially without any loss of signal in the diagnostic assay. Furthermore, we found there was no significant difference in assay readout whether the positive sample was loaded first or last in a series of 20 samples. These results demonstrate that different pooling strategies can lead to increased process efficiencies for COVID-19 clinical diagnostic testing.

## Introduction

The novel SARS-CoV-2 virus first emerged in Wuhan, China in January 2020 and has since rapidly spread around the world at an unprecedented scale. (1-3) The resulting COVID-19 global pandemic is challenging the existing supply chain for the diagnostic testing pipeline, leading to intermittent shortages in key supplies that have created testing bottlenecks. Diagnostic testing to identify individuals infected with SARS-CoV-2 plays a key role in controlling the COVID-19 pandemic by providing actionable information for quarantine strategies and population-level disease surveillance. In areas where the prevalence of COVID-19 cases is relatively low, one strategy to alleviate supply chain issues is to perform sample pooling, in which multiple clinical samples are combined and tested as a pooled batch. The pool can be categorized as negative with a single analysis if all the individual samples within the pool are negative for SARS-CoV-2, thereby eliminating the need to analyze every sample individually. This pooled testing strategy has been previously used to screen for sexually-transmitted diseases and to test blood banks for Hepatitis B and C, Zika virus and HIV. (4-5) Given that supply chain issues may still prove to be an obstacle to establishing comprehensive testing regimens as the need for testing continues to ramp up, pooling strategies will serve to conserve scarce resources and labor.

Several labs have tested various pooling strategies with SARS CoV-2 samples to examine such parameters as varying pool size versus loss of assay sensitivity and increase in false negatives. (6-8) Computer simulations and mathematical modeling have also been performed to determine optimal pooling techniques. (9) The US Food and Drug Administration (FDA) has granted an Emergency Use Authorization to Quest Diagnostics to authorize its SARS-CoV-2 qRT-PCR test for use with pooled samples containing up to 4 individual swab specimens. (10) The Quest test is the first COVID-19 diagnostic test to be authorized for use with pooled samples. Here, we have evaluated two different strategies for pooling independent nasopharyngeal (NP) swab samples: (1) a Dilution study to assess qRT-PCR assay efficiency using a range of viral load NP samples in pools up to 30, and (2) a Reloading study to assess the efficacy of reloading QIAamp viral RNA extraction columns numerous times using a single positive sample and multiple negative samples. We expect that these pooling results can help to alleviate the strain on the supply chain of diagnostic testing supplies and ultimately lead to reduced disease transmission. (11)

## Materials and Methods

### Samples and Assay

Known positive and known negative clinical nasopharyngeal swab samples were obtained from either the New Mexico Department of Health Scientific Laboratory Division (NM DOH SLD) or an internal pilot study approved by the IRB. Viral RNA was extracted from clinical samples using the QIAamp Viral RNA Mini Kit (Qiagen). For RT-qPCR analysis, we used EUA-approved TaqPath 1-Step RT-qPCR Master Mix (ThermoFisher) or qScript XLT 1-Step RT-PCR kit (Quantabio), with SARS-CoV-2 primers and probes from IDT (2019-nCov_N1, 2019-nCov_N2, Human RNase P), and the ABI 7500 Fast Dx instrument. As stipulated in the 'CDC 2019-nCoV Real-Time RT-PCR Diagnostic Panel', a human specimen control (HSC) was included with each batch of RNA extractions as an internal control and processed in parallel with the test conditions for all experiments.

### Dilution Study

In the first dilution experiment, a separate pool of negative diluent was made for each dilution ratio (1:5, 1:10, 1:20, and 1:30) using independent negative samples (i.e. 4, 9, 19, and 29 samples respectively). Each diluent pool was combined with 4 independent positive samples. In the second dilution experiment, one large pool of negative diluent was made from 29 independent negative samples and used across all dilution ratios, with 4 independent positive samples in each ratio. Sample pools for both experiments were prepared in 4 dilution ratios as follows: (1) 1:5 dilution = 30μl positive sample + 120μl pooled negative diluent, (2) 1:10 dilution = 20μl positive sample + 180μl pooled negative diluent, (3) 1:20 dilution = 10μl positive sample + 190μl pooled negative diluent, and (4) 1:30 dilution = 10μl positive sample + 290μl pooled negative diluent. A final volume of 140μl was then taken from each condition and processed for viral RNA extraction as per the QIAamp kit instructions. RT-qPCR was performed using TaqPath 1-Step RT-qPCR Master Mix (ThermoFisher). Data are shown in Table 1 and Table 2.

**Table 1.** Ct values (for N1, N2, and human RP) from dilution conditions (1:5, 1:10, 1:20, and 1:30) made using the first set of 4 independent positive clinical samples (A, B, C, and D). Undiluted positive samples are included as baseline controls.

| Condition | Ct N1 | Ct N2 | Ct RP |
| --- | --- | --- | --- |
| Undiluted Pos A | 28.58 | 28.75 | 30.59 |
| Pos A Dilution 1:5 | 31.01 | 32.42 | 29.39 |
| Pos A Dilution 1:10 | 31.89 | 32.66 | 29.56 |
| Pos A Dilution 1:20 | 32.96 | 34.50 | 31.46 |
| Pos A Dilution 1:30 | 33.14 | 33.63 | 29.57 |
| Undiluted Pos B | 29.08 | 30.25 | 27.03 |
| Pos B Dilution 1:5 | 33.00 | 34.28 | 29.26 |
| Pos B Dilution 1:10 | 34.49 | 36.50 | 29.30 |
| Pos B Dilution 1:20 | 35.36 | 37.70 | 31.17 |
| Pos B Dilution 1:30 | 36.31 | 37.86 | 29.56 |
| Undiluted Pos C | 35.38 | 36.23 | 33.17 |
| Pos C Dilution 1:5 | 38.10 | 38.61 | 29.49 |
| Pos C Dilution 1:10 | 35.83 | 38.66 | 29.55 |
| Pos C Dilution 1:20 | N/A | 38.15 | 31.36 |
| Pos C Dilution 1:30 | N/A | N/A | 29.77 |
| Undiluted Pos D | 36.85 | 36.79 | 31.57 |
| Pos D Dilution 1:5 | 35.97 | 38.02 | 29.55 |
| Pos D Dilution 1:10 | 37.05 | N/A | 29.44 |
| Pos D Dilution 1:20 | 37.37 | 39.02 | 31.68 |
| Pos D Dilution 1:30 | N/A | N/A | 29.52 |

Values shown are an average of two replicate RT-qPCR measurements. Red values indicate that one of the two measurements was Undetermined. N/A indicates that both measurements were Undetermined.

**Table 2.** Ct values (for N1, N2, and human RP) from dilution conditions (1:5, 1:10, 1:20, and 1:30) made using the second set of 4 independent positive clinical samples (E, F, G, and H). Undiluted positive samples are included as baseline controls.

| Condition | Ct N1 | Ct N2 | Ct RP |
| --- | --- | --- | --- |
| Undiluted Pos E | 31.57 | 33.30 | 29.97 |
| Pos E Dilution 1:5 | 34.48 | 35.87 | 33.26 |
| Pos E Dilution 1:10 | 35.50 | 37.42 | 33.38 |
| Pos E Dilution 1:20 | 35.37 | 39.83 | 34.83 |
| Pos E Dilution 1:30 | 37.49 | N/A | 35.26 |
| Undiluted Pos F | 34.33 | 35.77 | 33.25 |
| Pos F Dilution 1:5 | 37.45 | 38.08 | 35.80 |
| Pos F Dilution 1:10 | N/A | N/A | 36.51 |
| Pos F Dilution 1:20 | 38.03 | 39.70 | N/A |
| Pos F Dilution 1:30 | N/A | N/A | 37.7 |
| Undiluted Pos G | 28.55 | 30.26 | 29.26 |
| Pos G Dilution 1:5 | 31.43 | 33.12 | 31.09 |
| Pos G Dilution 1:10 | 32.66 | 34.17 | 32.32 |
| Pos G Dilution 1:20 | 34.36 | 35.63 | 32.92 |
| Pos G Dilution 1:30 | 33.88 | 36.45 | 33.61 |
| Undiluted Pos H | 24.83 | 26.65 | 26.18 |
| Pos H Dilution 1:5 | 27.66 | 29.67 | 28.63 |
| Pos H Dilution 1:10 | 28.80 | 30.87 | 30.13 |
| Pos H Dilution 1:20 | 29.98 | 31.79 | 30.93 |
| Pos H Dilution 1:30 | 30.32 | 32.66 | 31.41 |

Values shown are an average of two replicate RT-qPCR measurements. Red values indicate that one of the two measurements was Undetermined. N/A indicates that both measurements were Undetermined.

### Reloading Study

QIAamp viral RNA extraction columns were reloaded with 5, 10, or 20 separate samples (i.e. 5x, 10x, or 20x), using 1 positive sample along with 4, 9, or 19 independent negative samples respectively (140μl each). For each sample, 140μl was combined with 560μl Buffer AVL (with carrier RNA) + 560μl ethanol prior to column loading, as per the QIAamp kit instructions. For each series, the entire extraction volume was passed through one column in sequential centrifugation steps, loading 630μl each time. As such, 10, 20, or 40 centrifugation steps were needed to load 5, 10, or 20 samples respectively. In each series, the positive sample was loaded onto the column either as the first or last sample. RT-qPCR was performed using TaqPath 1-Step RT-qPCR Master Mix (ThermoFisher) for the experiment shown in Table 3, and qScript XLT 1-Step RT-PCR kit (Quantabio) for the experiment shown in Table 4.

**Table 3.** Ct values (for N1, N2, and human RP) from RNA extraction column reloading conditions (5x, 10x) using one positive sample loaded either first or last in each series. The positive sample with no reloads is included as a baseline control.

| Condition | Ct N1 | Ct N2 | Ct RP |
| --- | --- | --- | --- |
| Pos No Reloads | 34.39 | 36.84 | 33.36 |
| 5x series (Pos First) | 34.05 | 34.64 | 32.14 |
| 10x series (Pos First) | 34.77 | 35.77 | 32.65 |
| 5x series (Pos Last) | 34.12 | 35.07 | 32.33 |
| 10x series (Pos Last) | 34.41 | 35.54 | 32.29 |

Values shown are an average of two replicate RT-qPCR measurements.

**Table 4.**
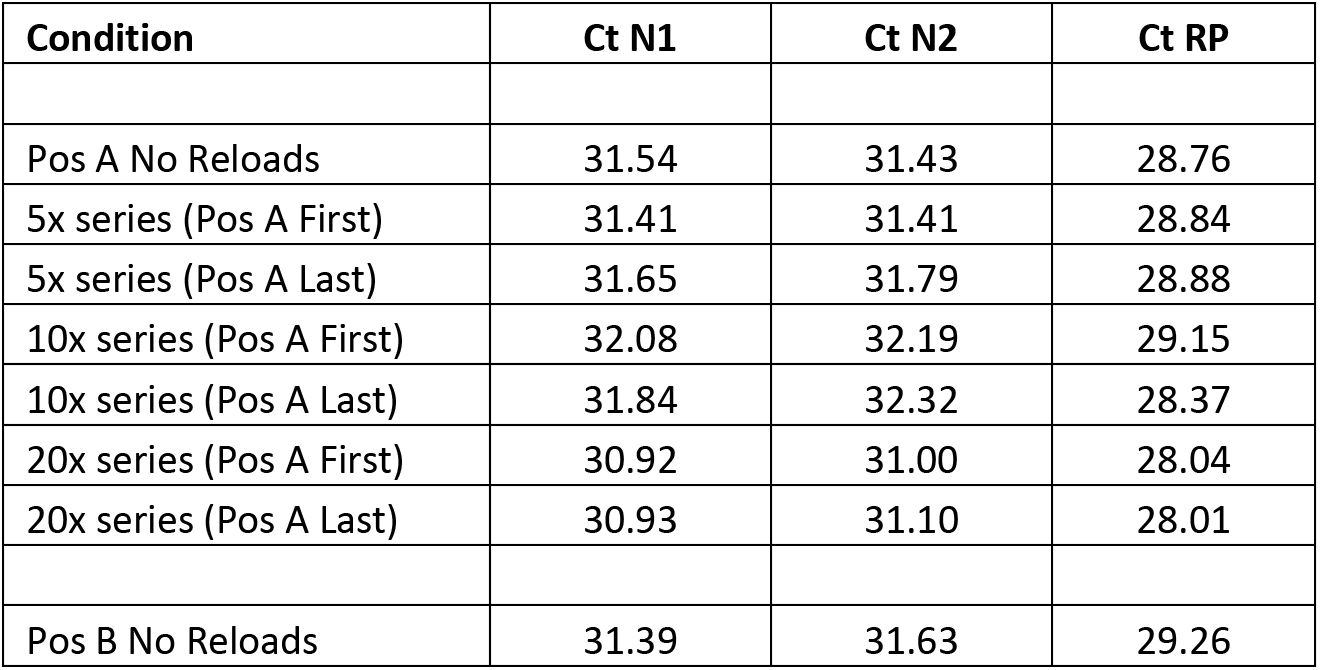
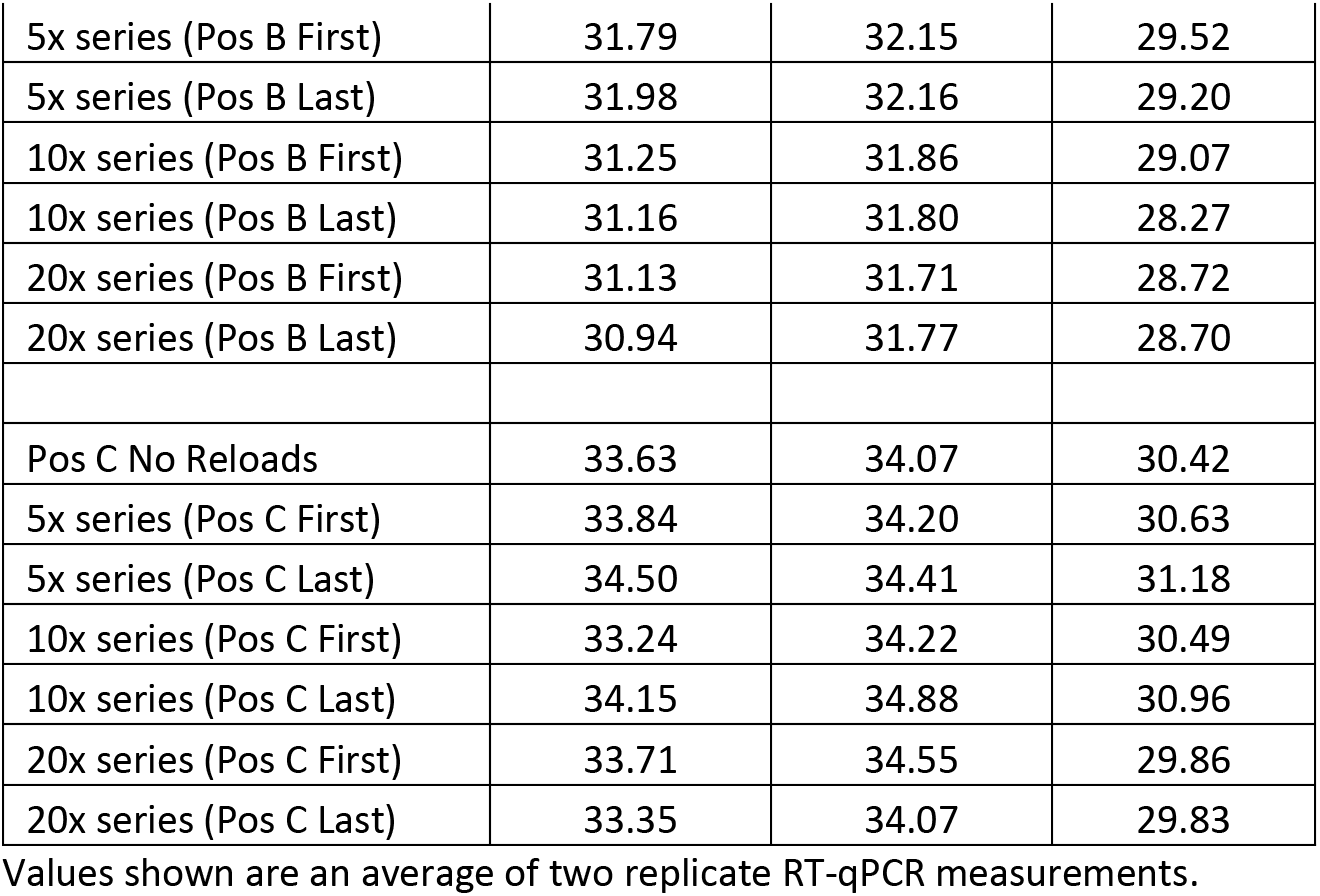
Ct values (for N1, N2, and human RP) from RNA extraction column reloading conditions (5x, 10x, 20x) using one positive sample loaded either first or last in each series. Positive samples with no reloads are included as baseline controls.

## Results and Discussion

### Dilution Study

To determine the lowest viral load (i.e. the highest Ct value) that can still be detected as a positive in a sample pool, we performed our dilution study with positive clinical samples that exhibit relatively high Ct values (>28). We created 4 dilution ratios of positive to negative samples (1:5, 1:10, 1:20, and 1:30) using 4 independent positive samples per experiment. Viral RNA was extracted from each condition, then RT-qPCR was performed to assess the diagnostic assay's sensitivity to the positive sample within each pool, and to determine the dilution ratio at which we would observe false negative readings.

For positive sample A (Pos A), with an undiluted Ct value of ~29 for both N1 and N2, we observed that high Ct values were detectable even at the 1:30 dilution. (Table 1) However, as the undiluted Ct value of the positive samples increased (Pos B, Pos C, and Pos D), we observed an increase in false negative readings. For Pos B, with an undiluted Ct value of ~29 for N1 and ~30 for N2, we found that one of the RT-qPCR measurements in the 1:20 pool was undetectable. For Pos C and Pos D, with undiluted Ct values >35 for both N1 and N2, there was an increased loss of assay signal in the 1:20 and 1:30 dilution ratios.

In the second dilution experiment, we tested 4 additional positive samples of varying Ct value ranges and observed similar results (Table 2). For Pos G and Pos H, with undiluted Ct values of ~29 and ~25 for N1 and ~30 and ~27 for N2, respectively, we detected reliable RT-qPCR signal even at the 1:30 dilutions. For Pos E, with undiluted Ct values of ~32 for N1 and ~33 for N2, we found the assay measurement for N2 to be undetectable at the 1:30 dilution. For Pos F, with undiluted Ct values of ~34 for N1 and ~36 for N2, we observed false negative readings starting at the 1:10 dilution. Our results suggest that positive clinical samples with Ct values at ~30 can be reliably detected from within a pool of up to 30 samples in which the other 29 are negative. Positive clinical samples with Ct values at ~35 can still be detected at a 1:5 dilution, i.e. in a pool with 4 other negative samples.

### Reloading study

There were two objectives in the reloading study: (1) to test the QIAamp viral RNA extraction column's ability to be reloaded effectively numerous times with multiple clinical samples as a means to pool samples without dilution, and (2) to determine if there is a difference in viral RNA output, as detectable by RT-qPCR, when the positive sample is loaded first compared to when the positive sample is loaded last in a series of 5, 10, or 20 samples.

In the first reloading experiment, 1 positive sample (Ct >33) and either 4 or 9 negative samples (for 5x or 10x series respectively) were prepared for viral RNA extraction as per the QIAamp kit instructions. We used a positive sample with a low viral load to test whether reloading an extraction column leads to a loss of signal in the RT-qPCR diagnostic assay or the occurrence of false negative results. Instead of using a separate column for each sample at the column-loading step, samples in the 5x and 10x series were loaded sequentially onto one column using repeated centrifugation steps, with the positive sample loaded either first or last in each series. Each column was subsequently processed and analyzed as one RNA sample. Table 3 shows a negligible loss of assay signal from reloaded columns under all conditions tested, indicating that RNA extraction efficiency was unaffected by the multiple reloading steps. All Ct values are comparable to the positive sample with no reloads (Ct value of ~34 for N1).

In the second reloading experiment, we tested 3 additional positive samples (all with undiluted Ct values >30) and included an additional reloading condition (20x series: 1 positive and 19 negatives). Even in the 20x series, where a single column was reloaded 40 times, we again observed a negligible loss of assay signal from reloaded columns under all conditions tested. (Table 4)

These results indicate that up to 20 clinical samples can be loaded onto a single extraction column without any significant loss of signal in the downstream RT-qPCR assay. The position of the positive sample in the reloading series (either first or last) also did not make a significant difference to RNA output, as determined by assay signal. We do note that reloading extraction columns using multiple centrifugation steps is time-consuming, adding 2 hours to the processing time needed for 20 samples. Use of a vacuum manifold to replace the centrifugation steps could reduce the additional time needed to load samples sequentially. Furthermore, reloading as a pooling strategy could be incorporated into the design of automated processes using existing liquid handling robotics platforms to greatly increase sample throughput for COVID-19 diagnostic testing.

Taken together, our results demonstrate that clinical sample pooling strategies can lead to increased process efficiencies for COVID-19 diagnostic testing in areas where the incidence of infection is relatively low. Thus pooling will help to alleviate the strain on healthcare services and may prove particularly useful in informing policymakers for targeted populations, such as return-to-work or school, to reduce disease transmission.

## Data Availability

N/A

## Acknowledgements

We thank Dr. Michael Edwards at the New Mexico Department of Health Scientific Laboratory Division (NM DOH SLD) for the COVID-19 clinical samples. Research was supported by the DOE Office of Science through the National Virtual Biotechnology Laboratory, a consortium of DOE national laboratories focused on response to COVID-19, with funding provided by the Coronavirus CARES Act.

